# Covid19data.website

**DOI:** 10.1101/2020.08.14.20174995

**Authors:** Dhafer Malouche

**Author notes:** Corresponding author;, website: https://dhafermalouche.net.

## Abstract

Covid19data.website Project is a website that contains more than 250 Dashboards about the COVID-19 Toll areas affected by the virus. You can follow-up every day using this website the number of COVID-19 cases, deaths, active cases, the shares by 1 Million population, the mortality rate, the active rate in every country, continent, and territory and all States in the United States. Unlike the other websites, you can also follow-up an estimation of the reproduction number in the previous sixteen days. These statistics tell you how many secondary infections are likely to occur in a specific area.

Furthermore, I provide a classification algorithm of the countries and the affected areas. It’s based on the observation during the previous 14 days of six criteria. A global score is then computed, allowing to evaluate the COVID-19 safeness toll of each area.

## 1. INTRODUCTION

The first COVID-19 positive case was confirmed in China on December 1st, 2019, according to the data available in the R package in Github implemented in [1]. More than 200 days later, this situation is then transformed into a worldwide outbreak affecting all the countries. According to the WHO^1^, the number of detected COVID-19 positive reached in August 13th, more than 20 million cases in the World. The United States is leading the countries with more than 5 million cases, with more than 160,000 deaths.

Many organizations have deployed web site where they keep the people always informed about the COVID-19 toll in the World and the countries affected by this virus. WHO has then published a dynamic website^2^ containing a dynamic and an explorer tab-set where you can explore the COVID-19 toll in each country and territories in the World. We can visualize the global map of the number of confirmed cases, the number of deaths, the shares by 1 million population of the number of cases, and the number of deaths. We can also observe the time series of these statistics by territories and countries.

On the web, you can find several projects providing a lot of statistics about the COVID-19 outbreak. I have recently found a very friendly interactive website^3^ designed by Avi Schiffmann, a high schooler from Washington State, USA. This website contains data tables displaying the Covid19 toll for each country. It shows the number of COVID-19 tests in each country. On this website, there’s a global map where the COVID-19 statistics pop up when we click on the country name. The available statistics are again the number of cases, deaths, active cases, and deceased cases.

Another popular website deployed because of the pandemic situation was the John Hopkins University website^4^. It contains a lot of data resources and visualizations: Maps, charts, articles, and data.

I had then started creating a website about COVID-19 statistics. I wanted to create a website where a user can learn more about the COVID-19 outbreak. Besides the standard statistics like the number of confirmed cases, the number of deaths, the number of recovered, the number of cases by 1 Million population and others, I provide in this website two crucial information that can help anyone to have a better follow up of the pandemic: the reproduction number (see [2, 3]) and a country COVID-19 classification based on six criteria.

The data used in this website comes from the R package named COVID19^5^. This package provides a comprehensive fine-grained case data, merged with exogenous variables (see [5]). I also use a second R package nCov19 to check the previous data and make some edits on the mistakes when it’s needed (see [1]).

This website is updated every 24 hours. It’s build using R Studio and flexdashboard package.

Most of the visualization are performed using dygraph and highcharter packages. They are interactive graphs with pop-ups that can help the users to read the data better.

The following section will explain the several features existing on the website in more detail, and I will show how the COVID-19 classification algorithm works.

## 2. WEBSITE DESCRIPTION

This section is composed of three subsections. In the next one, I will explain the COVID-19 indicators that are displayed on the website. In the second subsection, I describe the front page of covid19data.website and the architecture of my website. The third section is devoted to the classification algorithm.

### A. Covid19 indicators

The data used in this website provides mainly three statistics that are available for each country *w*, affected by the COVID-19, and at the day *t*:

- *C*(*t, w*) is the number of cumulative COVID-19 detected cases until the day *t* in the country *w* (see (1) in Figure fig-S1),
- *D*(*t, w*) is the number of deaths from COVID-19 until the day *t* in the country *w* (see (2) in Figure fig-S1),
- *R*(*t, w*) is the number of recovered cases until the day *t* in the country w (see (3) in Figure fig-S1).

From these statistics, I have computed many other indicators at the countries’ level, regions as defined by the World Bank, income regions, continents, sub-regions, and the aggregated statistics in the world. These indicators are defined as follows:

- the new cases (see (6) in Figure fig-S1):

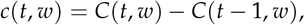
- the new death cases (see (7) in Figure fig-S1):

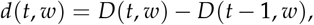
- the number of active cases (see (4) in Figure fig-S1):

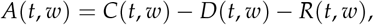
- known the population’s size *P*(*w*) in 2020 in a given country, region, and area *w*, we define by 1 Million population:
  - the total number of cases (see (9) in Figure fig-S1): 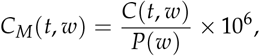
  - the total number of death cases (see (10) in Figure fig-S1): 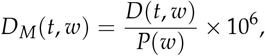
  - the total number of recovered cases (see (11) in Figure fig-S1): 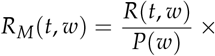
  - the total number of active cases (see (12) in Figure fig-S1): 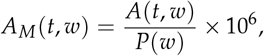
- the rate of
  - Mortality (see (5) in Figure fig-S1): 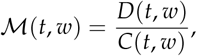
  - Active cases (see (8) in Figure fig-S1): 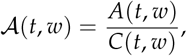
- the number of days to detect 50% of the total number of
  - cases (see (14) in Figure fig-S1):

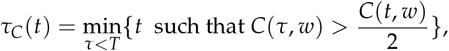
  - deaths (see (15) in Figure fig-S1):

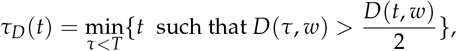
- during the previous 7 days, the number of
  - cases detected (see (20) in Figure fig-S1):

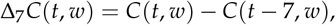
  - death cases (see (21) in Figure fig-S1)

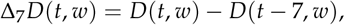
  - recovered cases (see (22) in Figure fig-S1)

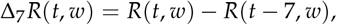
- the number of days since
  - the first detect COVID-19 positive case (see (16) in Figure fig-S1),
  - the first observed COVID-19 death ((see (19) in Figure fig-S1)),
- the reproduction number *R*(*t, w*), (see (13) in Figure fig-S1), estimated using the algorithm implemented in EpiEstim package implemented in *R* (see [2, 3]).

### B. Front page

The front page of covid19data.website is dedicated to the aggregated statistics in the World. It also contains two other indicators: the number of days to detect the last million cases and the number of days since the previous 100,000 deaths.

On the same front page, the user can also navigate to the other pages dedicated to lower-level aggregated statistics. From this page, the user can learn about the COVID-19 outbreak in

- the five continents: Africa, Asia, Europe, North, South America,
- Seven regions: defined in the World Bank Data Indicators website (see [6]),
- Four income region also defined in the World Bank Data Indicators (see [6]),
- The United States and the European Union.

Once the user goes to any of these region dashboards, he/she can learn about the COVID-19 toll of the countries belonging to them.

The same front page also contains three buttons that can be used to display three Tableau Dashboards built using the Software Tableau public^6^:

- Dashboard 1 (see (5) in Figure fig-S2): It’s an interactive scatter plot Statistics by 1 Million population versus the number of days. The user can use this Dashboard to visualize and compare, for example, the number of deaths by 1 million population, between the countries affected by the COVID-19. The user can also build figures of countries of a specific continent or Income region (see Figure fig-S3).
- Dashboard 2 (see (6) in Figure fig-S2) displays the rates of Mortality and Active cases (see Figure fig-S5).
- Dashboard 3 (see (7) in Figure fig-S2) displays a Global map colored according to my COVID-19 Safeness classification that will explain in detail below (see Figure fig-S4).

**Fig. S1.**
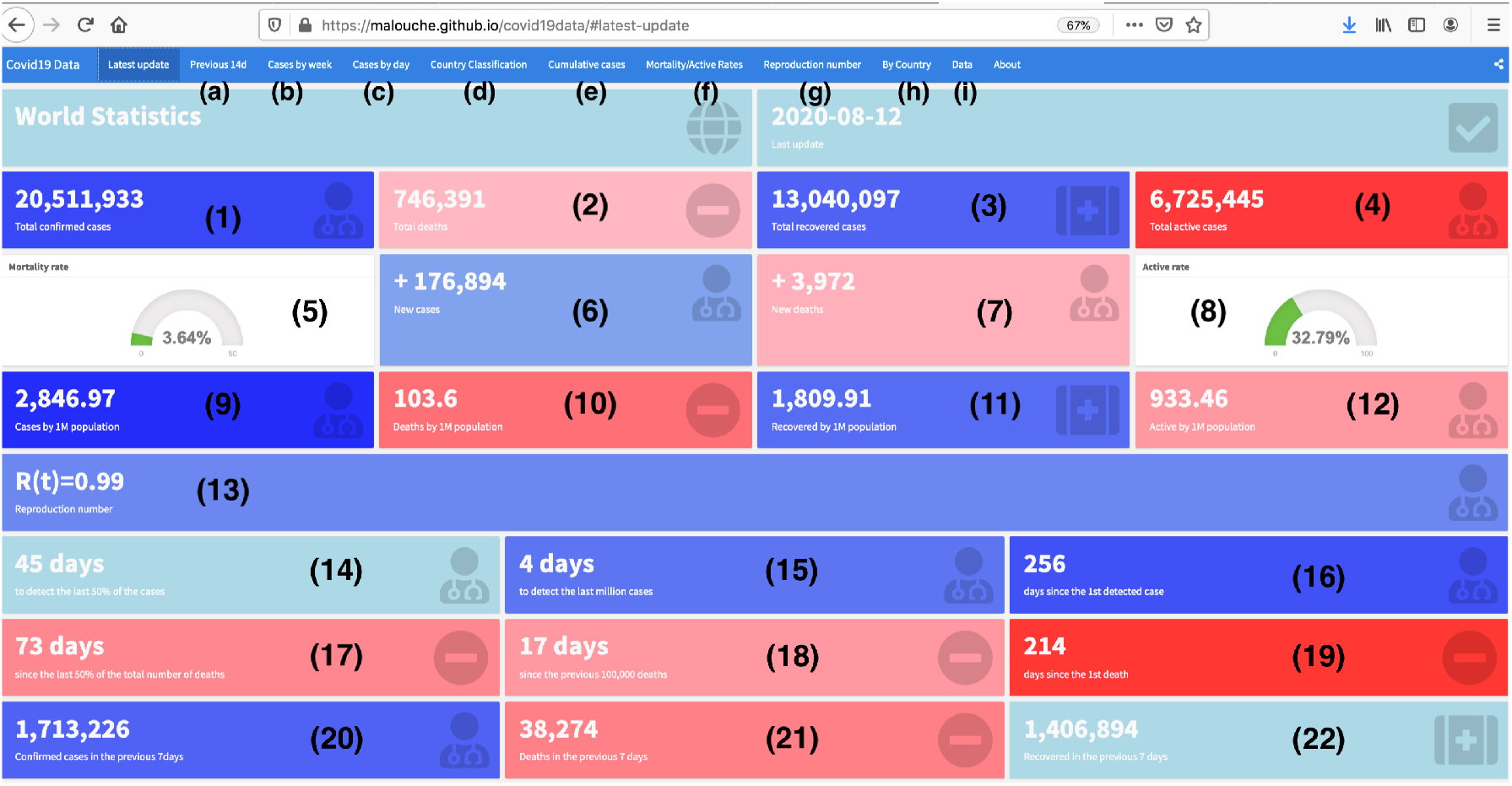
The top of the Front page of covid19data.website

On the top of the front page, you can also navigate to ten other pages:

- Previous 14d (see (a) in Figure fig-S1): it’s a page that displays time series in the previous 14 days of the cumulative cases, death cases of the continents, or the top 5 regions or the top 5 countries, or the five states according to each of these indicators.
- Cases by week (see (b) in Figure fig-S1): it displays also aggregated indicators by week. These figures are build using dygraph package in *R* (see [7]).
- Cases by day (see (c) in Figure fig-S1): it displays the time series of the indicators: *c*(*t, w*) the daily new cases and *d*(*t*, *w*) the daily new death cases. These figures are build using dygraph package in R (see [7]).
- Cumulative cases (see (e) in Figure fig-S1): it displays time series of the cumulative indicators *C*(*t, w*), *D*(*t, w*), *R*(*t*, *w*), and *A*(*t*, *w*), either together in the same graph or each one in a separate graph.
- Country Classification (see (d) in Figure fig-S1): It displays the result of my COVID-19 Safeness classification. It displays an interactive table build using DT package. I also display a short note explaining the detail of the classification algorithm.
- Mortality/Active rate (see (f) in Figure fig-S1): it displays the time series of the indicators *M*(*t*, w) and *A*(*t, w*).
- Reproduction number (see (g) in Figure fig-S1): it displays the time series of the reproduction number *R*(*t, w*) during the previous 60 days.
- By Country (see (h) in Figure fig-S1): It contains a button with the names of the countries sorted in alphabetical order. Each button will take you to the Dashboard of the associated country.
- Data (see (i) in Figure fig-S1): I display the whole data compiled from the packages COVID19 and nCov19. This data can be downloaded in several formats: CSV, XLS, TXT. You can also filter and sort the data according to any request.

**Fig. S2.**
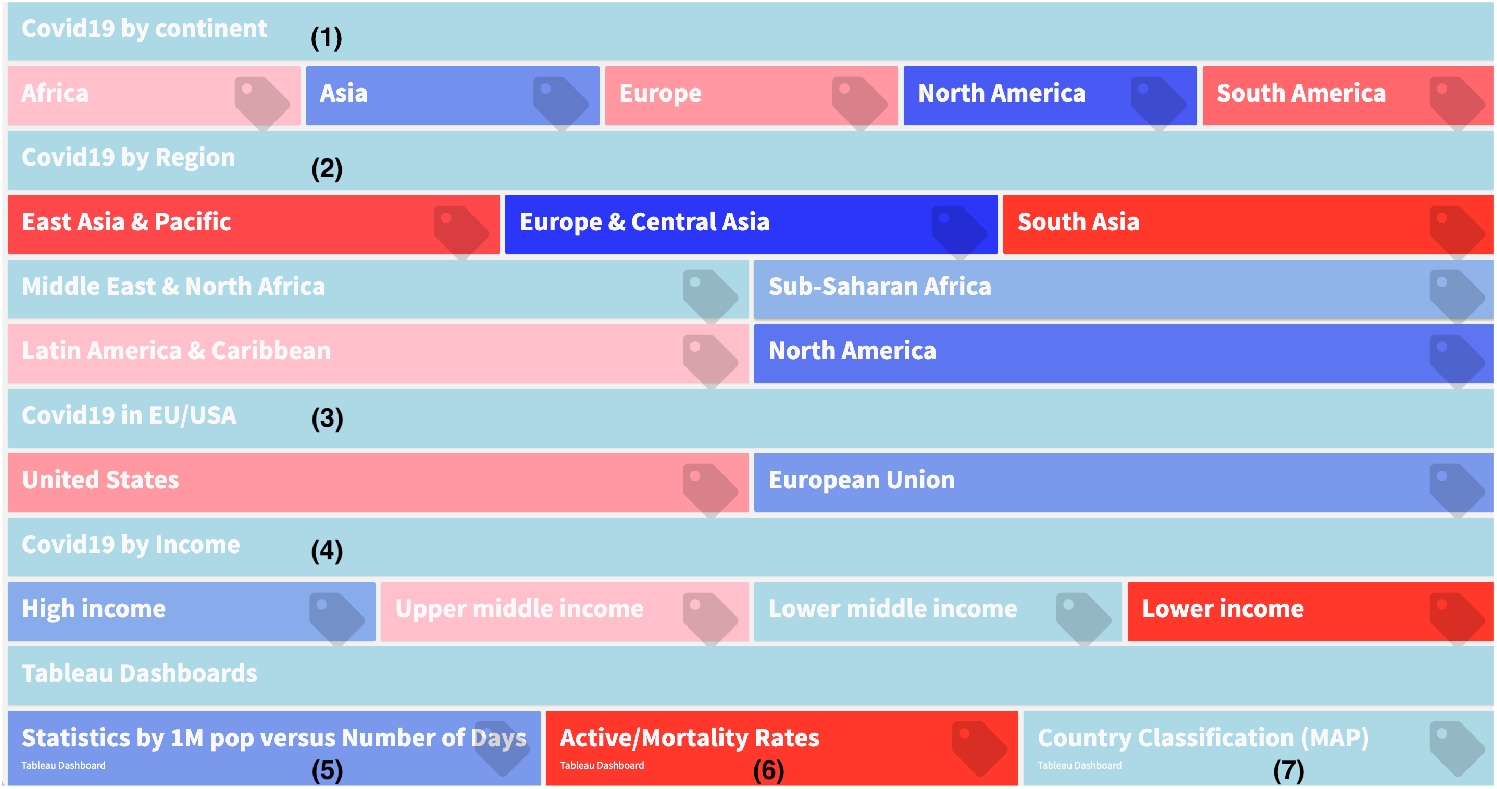
Second part of the Front page of covid19data.website

**Fig. S3.**
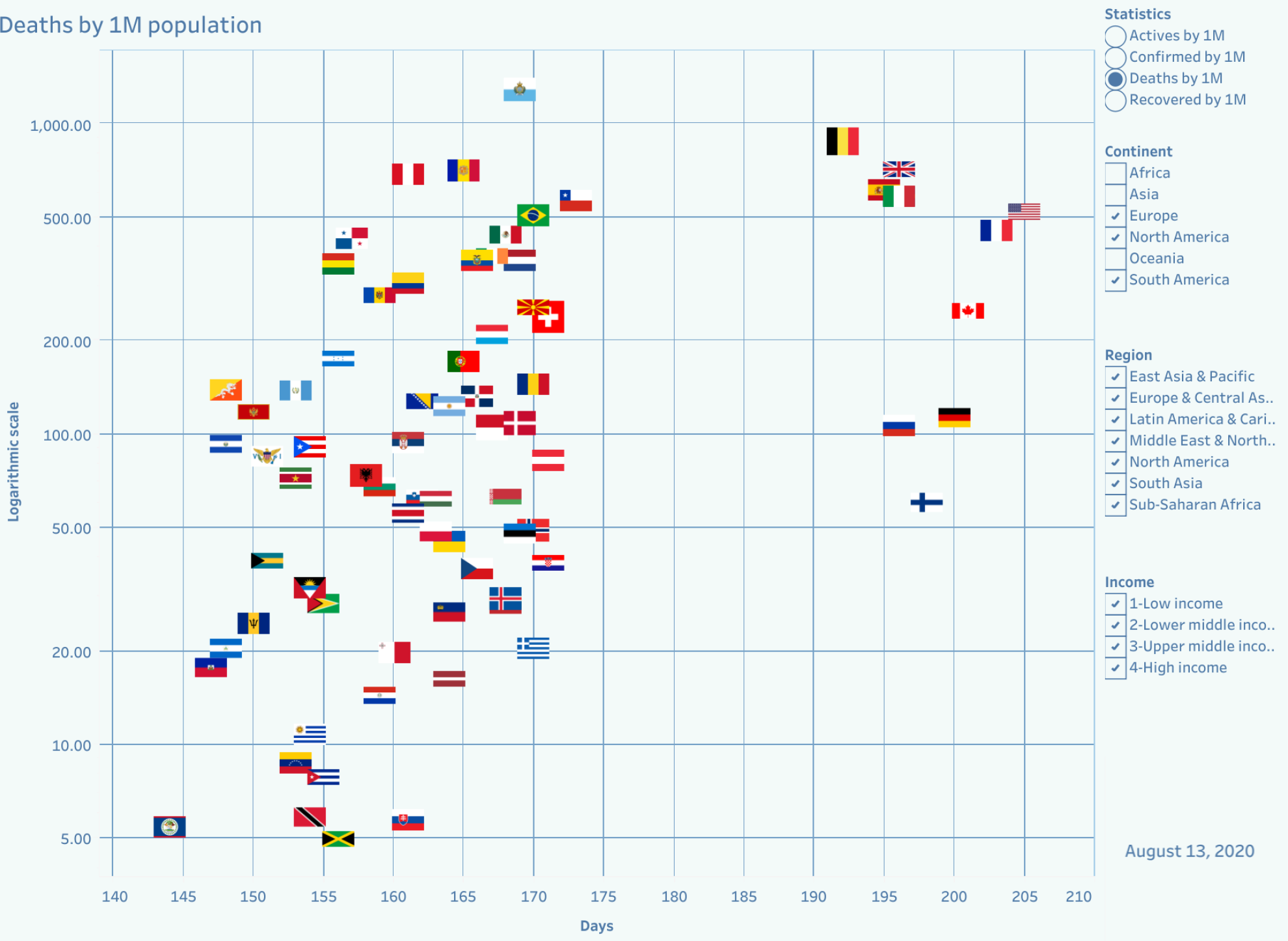
Statistics by 1 Million population Dashboard

**Fig. S4.**
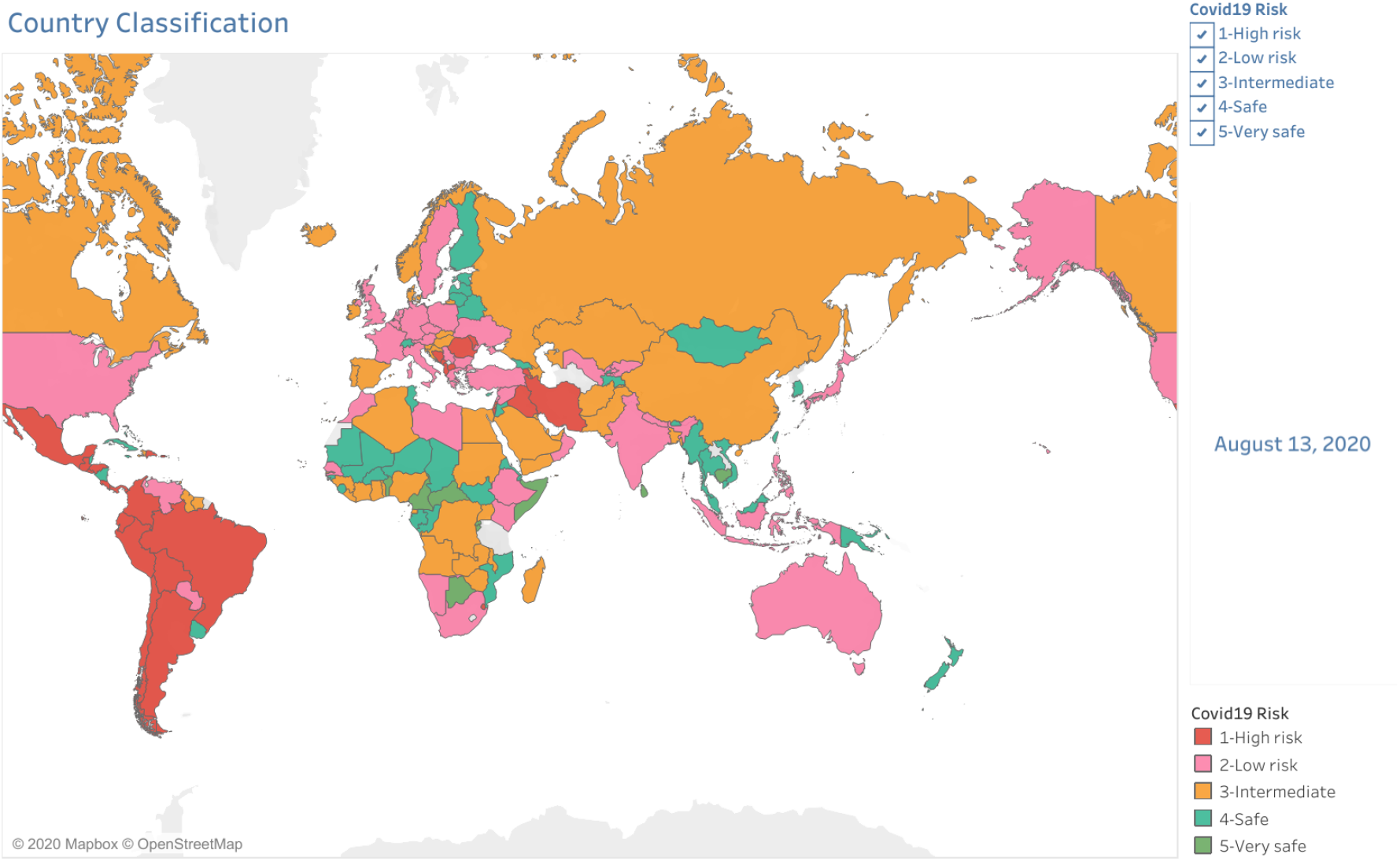
Country Classification Map Dashboard

**Fig. S5.**
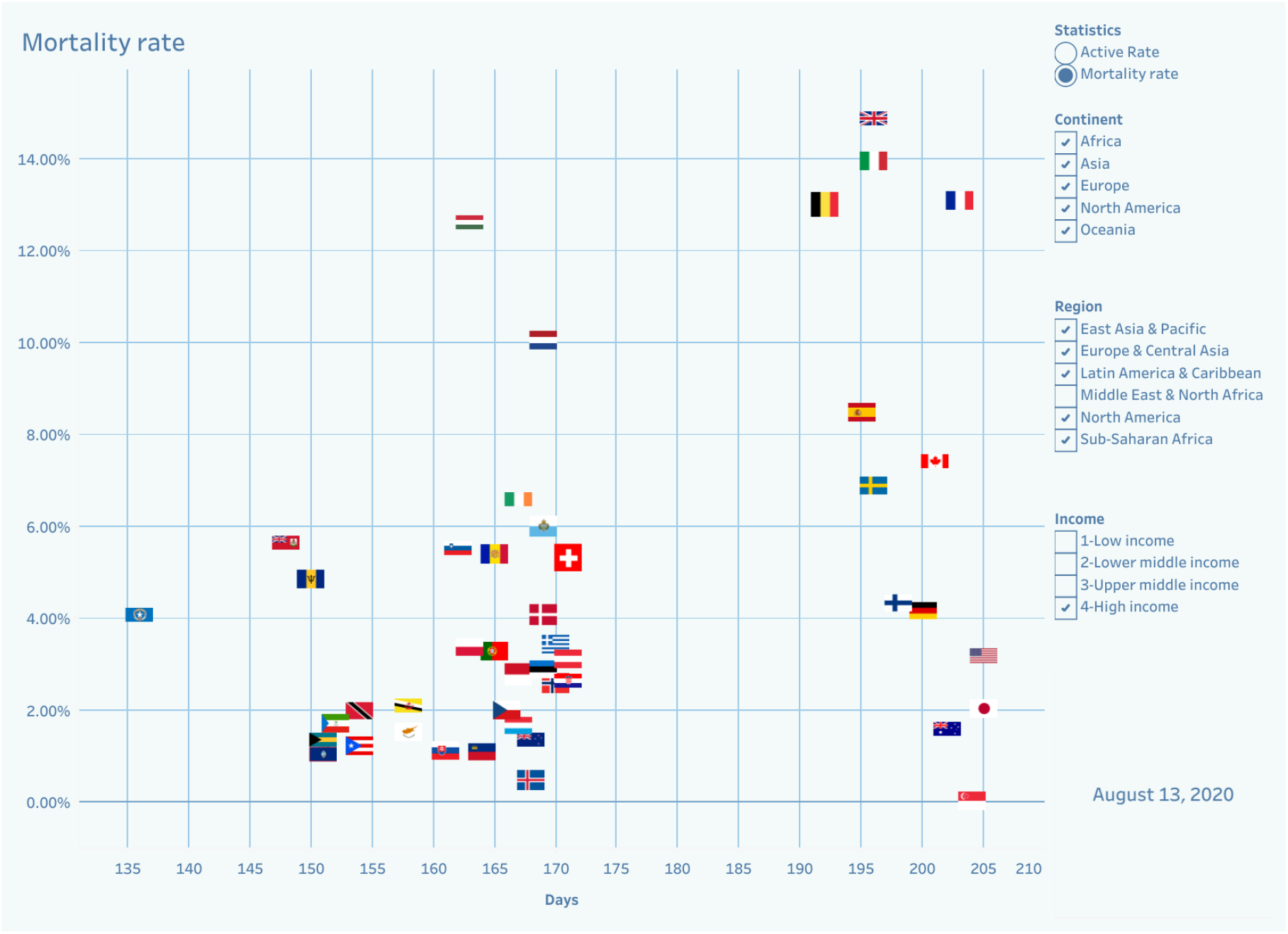
Mortality and Active rate Dashboard

### C. Covid19 Classification

One of the main contributions in covid19data.website is a country classification that helps to understand the COVID-19 toll in each country and provides a tool to make a comparison between countries. This classification algorithm is based on six criteria: the reproduction number, the growth of the active cases, the deaths, the total number of cases, and the deaths by 1 million population.

I consider first in this classification the following criteria:

4 indicators related to confirmed cases:

- C_1_ In the previous seven days, the upper band of the 95% CI of the reproduction number *R*(*t, w*) is lower than 1 in at least four days:

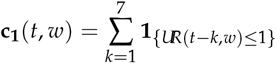

where 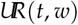 is the upper band of the the 95% CI of the *R*(*t, w*) at the day *t*, and the country *w*.

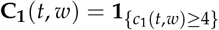
- C2 In the previous 14 days, the Active rate decreases at least in seven days:

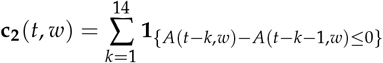

where *A*(*t*, *w*) is the number of active cases at the day t in the country *w*, and

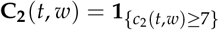
- C3 In the previous 14 days, the number of daily cases decreases in at least seven days:

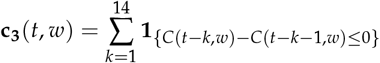

where *C*(*t, w*) is the number of daily cases at the day *t*, and

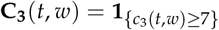
- C4 In the previous seven days, the number of cases by 1M population is lower than 21:

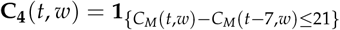

where *C_M_*(*t*) is the number of cases by 1M population at the day *t* in the country *w*.

And two indicators related to death cases:

- D_1_ In the previous 14 days, the number of daily deaths decreases in at least ten days:

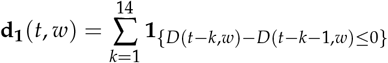

where *D*(*t, w*) is the number of death cases until the day *t*, and

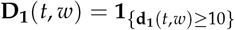
- *D*_2_ In the previous 14 days, the number of deaths by 1M population is lower than 14

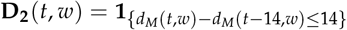

where *D_M_*(*t, w*) is the number of deaths by 1M population at the day *t* in the country

I compute then an Overall score as follows:

1. A first score at the day *t*:

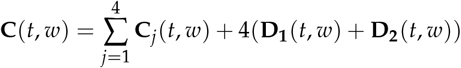 I give here a higher weight for the death indicators.
2. An overall score which is a weighted sum of the 1st score on the previous days:

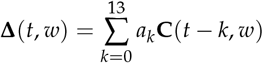

where 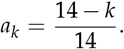 It’s easy to prove that *∆*(*t*) takes its values in [0,90]. I will consider then its standardized version ∆*_s_* (*t, w*) = ∆(*t, w*)/90 × 100 ∊ [0, 100%] as a percentage of the maximum score required.
3. From the standardized score ∆*_s_* (*t, w*) I create five categories from the highest risk to the very safe:

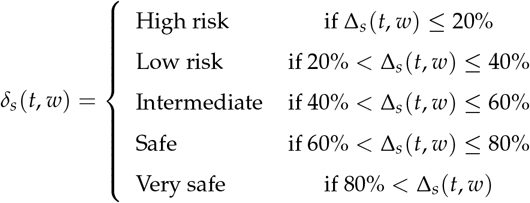

This country classification is then displayed by clicking on Country Classification (see (d) in Figure fig-S1) on the top of the front page, in the Covid19data.website, in an interactive table built with DT package. It allows you to filter, search, and download in several formats: CSV, XLS, PDF, and TXT. The user can then download a table with the classification scores and categories (see Figure fig-S6).

In each country/state Dashboard, the user can also learn about its safety score and category on the country/state front page. By clicking on the tab Covid19 classification, a time series of the previous 21 days of the country/state score is then displayed. The background of this chart corresponds to the colors associated with the classification category. It will then be easier for the user to understand the COVID-19 situation in the specific country/state and what the trend could in the next couple of days (see Figure fig-S7).

## 3. CONCLUSION

I have shown and explained in this paper how to use the Covid19data.website to learn about the COVID-19 outbreak in the World, in all the continents, in many regions, in all countries affected by the virus, and in the states of the United States. Using this website, you can learn about the reproduction number and the COVID-19 Safeness situation in all USA countries and states. Tableau Dashboards are also available on this website. They can help the user build his charts and compare the countries.

**Fig. S6.**
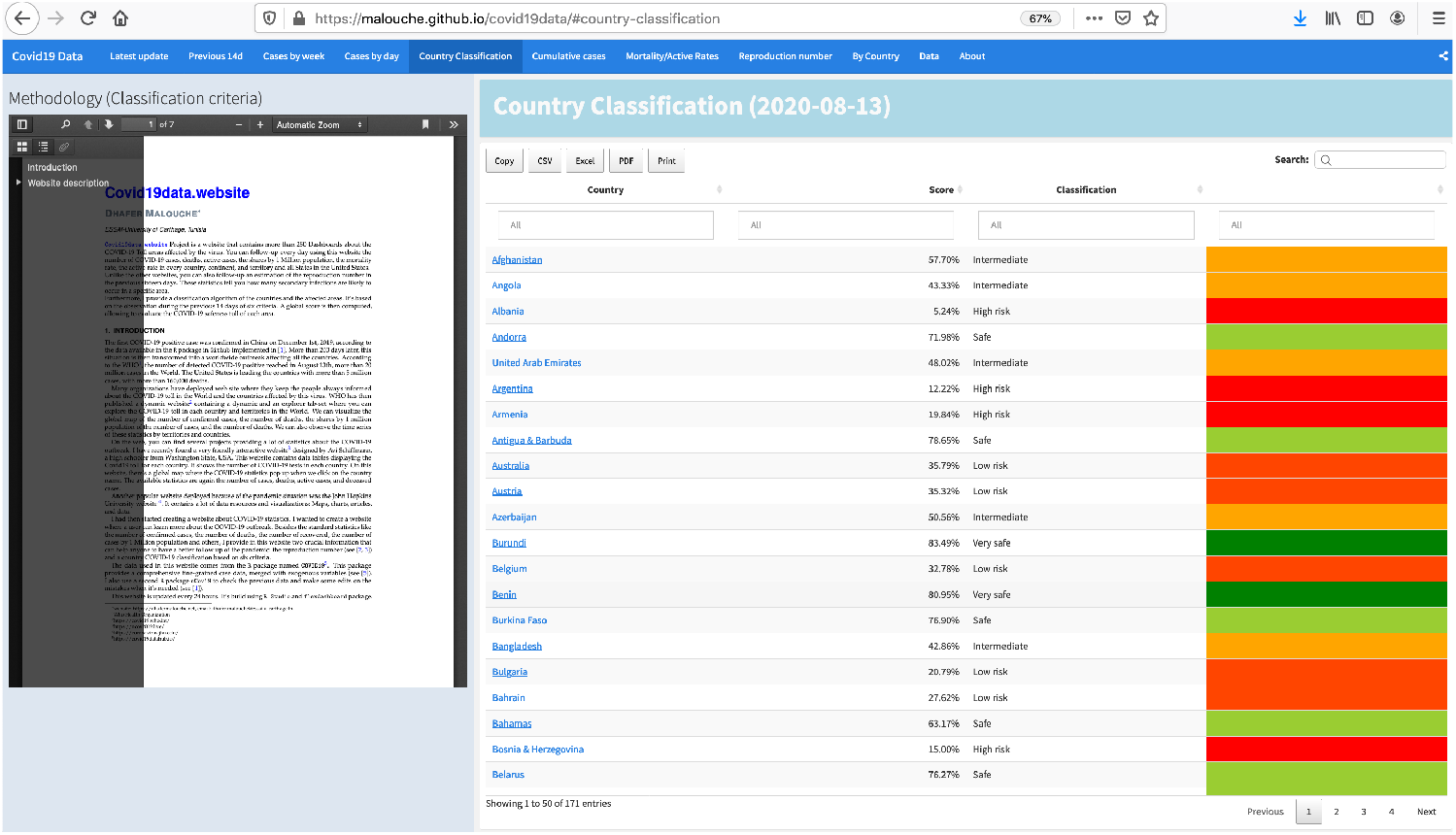
Table of the Country classification

**Fig. S7.**
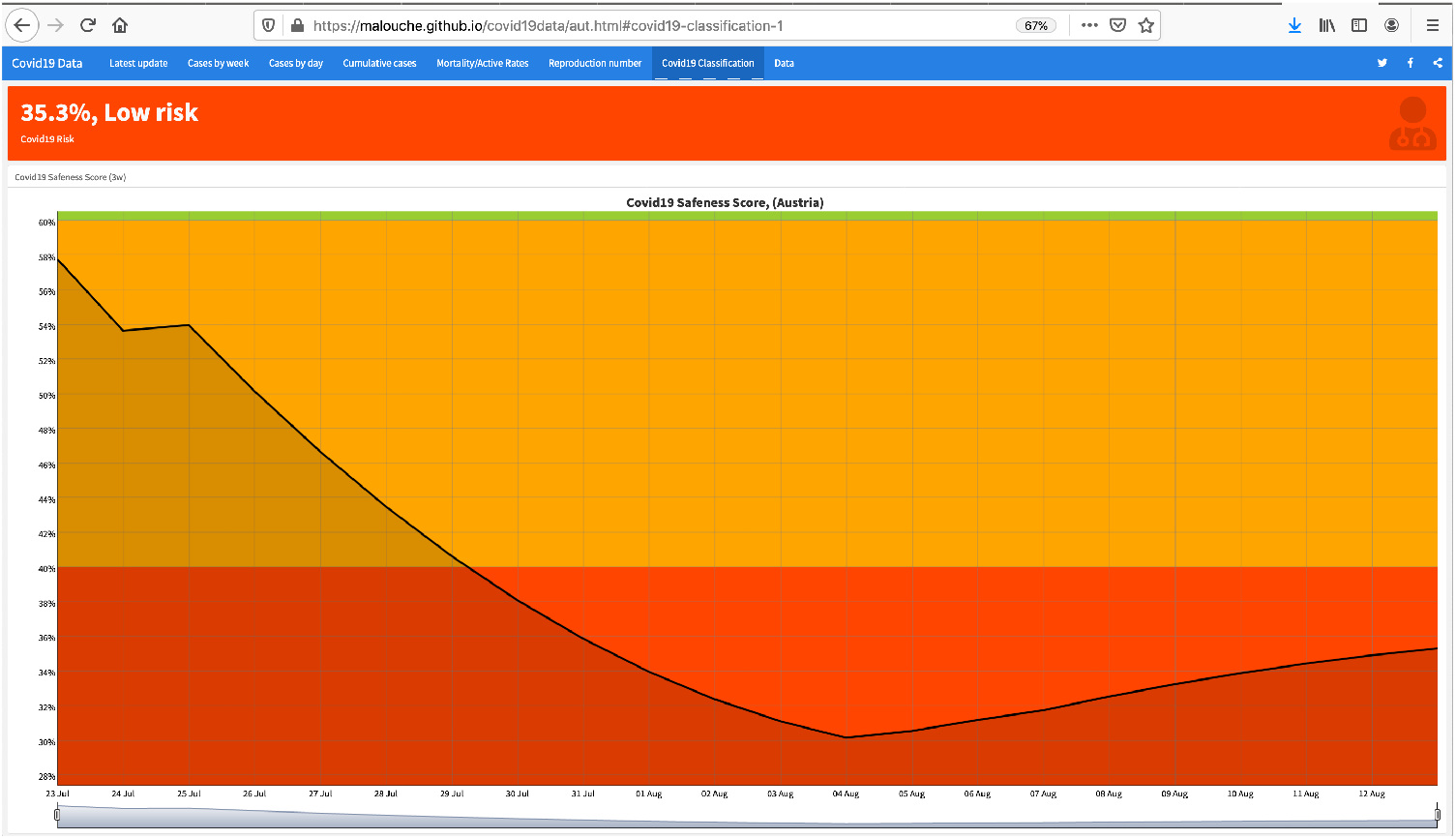
Time series of the country classification

## Data Availability

data extracted from COVID19 package in R software

https://covid19datahub.io

https://covid19data.website

## Footnotes

1 Who Health Organization

2 https://covid19.who.int/

3 https://ncov2019.live/

4 https://coronavirus.jhu.edu/

5 https://covid19datahub.io/

6 https://public.tableau.com/

